# Measurement of perceived healthcare discrimination and its implications on population health research: A systematic review protocol

**DOI:** 10.1101/2024.12.18.24319227

**Authors:** Michael D. Green, Ashleigh B. Harlow, Sarah E. Janek, Kevin P. Weinfert, Lesley Skalla, Emily C. O’Brien, M. Alan Brookhart, Ann Marie Navar, Perusi Muhigaba, Keisha L. Bentley-Edwards, Matthew E. Dupre, Roland J. Thorpe

## Abstract

**Purpose:** Discrimination in healthcare settings significantly impacts population health, affecting individuals of racial and ethnic minoritized identities, sexual and gender minoritized identities, and individuals with disabilities. Most measures of discrimination are derived from instruments assessing everyday discrimination experiences and do not adequately capture the specific nuances of healthcare-related discrimination. This systematic review protocol outlines a planned study that will evaluate and synthesize tools designed to measure discrimination in healthcare settings in the United States (U.S.). By providing a detailed overview of these measurement tools and their psychometric properties, this review will enhance understanding of healthcare discrimination and guide policy and practice towards improving health equity.

**Methods:** A comprehensive search will be conducted across four databases (PubMed, Embase, PsycINFO, and Web of Science) and supplemented by citation tracking of reviewed articles. The review will focus on studies involving measurements of perceived discrimination within healthcare settings in the United States. The primary outcomes include the reliability, validity, and specificity of discrimination measurement instruments. Quality assessment will use the COnsensus-based Standards for the selection of health Measurement INstruments (COSMIN) Risk of Bias tool. Our systematic review will be present findings based on the Preferred Reporting Items for Systematic Review and Meta-Analyses COnsensus-based Standards for the selection of Health Measurment INstruments of outcome measurements 2024 (PRISMA-COSMIN for OMIs 2024) reporting standard. The protocol is registered with PROSPERO (Registration number: CRD42024592267.

**Discussion:** This review will synthesize existing literature and offer a comprehensive evaluation of discrimination measurement tools to guide future health equity research. This review aims to contribute to the mitigation of healthcare inequities and the promotion of health equity by offering actionable insights into the measurement of discrimination in healthcare settings.

## Introduction

Discrimination within healthcare settings has profound implications on population health, influencing health outcomes across different groups in the United States (US), including racial and ethnic minoritized individuals, sexual and gender minoritized individuals, and older adults.[1-3] Discrimination, defined as the unjust or prejudicial treatment of individuals based on characteristics such as race, gender, or socioeconomic status, is typically measured as an experience in multiple settings through surveys and questionnaires. These tools assess perceptions of discrimination experiences across various life domains such as employment, education, and housing.[4] While these general measures are important for understanding the overall stressors individuals face, they do not capture the nuanced experience of discrimination within healthcare settings, where systemic racism is embedded in institutional policies and reinforced by the biases of individual clinicians.[5]

Discrimination in healthcare settings is connected to social stratification, a system which ranks and divides based on characteristics such as economic status, race, and education.[6-8] This results in unequal access to resources and services, manifesting in healthcare inequities in access and treatment quality. For instance, economically disadvantaged groups may face barriers to healthcare access, while racial and ethnic minoritized individuals may receive different standards of care due to systemic biases. Given these complexities, accurate measurement of discrimination within healthcare is crucial. Well validated measures enable researchers and policymakers to identify specific areas of inequity and develop targeted interventions. By identifying these tools, we can improve population health by addressing how discrimination manifests and subsequently impacts health outcomes, ultimately working toward more equitable healthcare systems.

Although systemic issues within healthcare are a crucial focus for population health researchers, accurately assessing individual experiences of discrimination remains equally important. Reports of patients not feeling empowered or heard illustrate the direct impacts of discrimination on health outcomes.[9] For instance, when patients feel ignored or disrespected by their healthcare providers, they may lose trust in the care they receive, making them less likely to follow medical advice or attend follow-up appointments. Similarly, poor communication during consultations can leave patients feeling confused about their diagnosis or treatment plan, which can further undermine their engagement in managing their health. Healthcare discrimination is particularly harmful because it directly affects clinical interpersonal interactions and subsequently health outcomes.[10, 11] Identifying healthcare discrimination, especially the settings and populations most affected, may allow for targeted interventions to combat it.

The existing evidence surrounding the clinical implications of non-context specific forms of discrimination is fragmented, with varying measurement tools leading to inconsistent results, hampering implementation of effective interventions by healthcare professionals and policymakers.[5, 12, 13] To date, there are no systematic reviews of measures for reporting discrimination in healthcare settings. There have been some literature reviews of the use of individual tools, without a focus on their measurement properties. For example, a recent literature review that examined the ‘Discrimination in Health Care Measure’ found it to be reliable across studies, as well as associated with adverse health outcomes, particularly in race- or ethnicity-based discrimination contexts.[14] This tool has been validated among African-American patients with only one study assessing its construct validity,[15] and only employed in small-scale, cross-sectional studies.[14] Additionally, it has been used without significant modifications across diverse populations and settings, with reliability generally assessed through Cronbach’s alpha, which has recognized constraints. Other studies have evaluated tools aimed at assessing healthcare treatment for targeted groups, such as a scale measuring medical mistrust among formerly incarcerated Black and Latino men,[16] and another assessing the humanization of medical care for transgender persons.[17] However, the overall clarity on perceived discrimination in healthcare as a construct, and the validity of tools designed to measure it, remains ambiguous overall population health. This review will focus on identifying the psychometric properties of these tools and synthesizing their use cases across different healthcare settings to better understand their applicability and effectiveness.

## Methods and Analysis

### Study Aim and Guidelines

We will conduct a systematic review of the existing literature which outline the properties (e.g. positive predictive value, negative predictive value, sensitivity, specificity, concordance statistic, and calibration) of measurement tools for patient-reported discrimination in healthcare settings. This systematic review will investigate how discrimination is being assessed across different population groups, highlighting the validity, strengths, and weaknesses of current measurement tools. This protocol describes the review methodology in accordance with the Preferred Reporting Items for Systematic Review and Meta-Analysis Protocols (PRISMA-P) guidelines (Supplementary Table 1).[18] The protocol is registered with the PROSPERO International Prospective Register of Systematic Reviews (Prospero ID CRD42024592267). Our review will be guided by the National Academies Press (formerly Institute of Medicine) systematic review standards, and then reported following the Preferred Reporting Items for Systematic Reviews and Meta-Analyses Consensus-based Standards for the Selection of Health Measurement Instruments for Outcome Measurement Instruments 2024 guidelines (PRISMA-COSMIN for OMIs 2024).[19, 20] These guidelines contain 54 components which should be identified and reported when reviewing tools. It emphasizes transparency in open science practices, including funding disclosures, registration, protocol accessibility, and competing interests. Additionally, the PRISMA-COSMIN for OMIs reporting standards specifies requirements for detailing eligibility criteria, risk of bias assessment, synthesis methods, and reporting results.

### Eligibility criteria

An overview of our inclusion/exclusion criteria is presented in Table 1. To be included, studies must involve development and/or evaluation of discrimination measurement for humans in healthcare settings, be original empirical research, and be published in peer-reviewed journals in English. Studies will be excluded if they are not focused on focused on patient- or caregiver-reported discrimination, nor attempting to ascertain or capture discrimination. Publications will also be excluded if they are review articles, study protocols, pre-prints, concept analysis papers, commentaries, case reports, or expert opinions. Studies focusing on discrimination outside of the United States are also excluded. We recognize that restricting our review to studies published in English and conducted within the United States limits the global applicability of our findings. Due to the unique sociohistorical context of the U.S. healthcare system, our team will be unable to appropriately synthesize findings from different countries, as discrimination may manifest differently within their healthcare policies and systems. While we will include non-English manuscripts in the initial review, full translations will only be performed if they pass the screening stage. These limitations will be addressed in the final systematic review manuscript.

**Table 1.** Inclusion/Exclusion Criteria for Study Selection Process.

| **Population,**<br>**Intervention,**<br>**Comparison,**<br>**Outcomes, Study**<br>**Design (PICOS)** | **Inclusion** | **Exclusion** |
| --- | --- | --- |
| **Population** | Studies involving human patients in healthcare settings within the U.S., focusing on the development or evaluation of discrimination measures. | Studies conducted outside of the U.S. or not involving patient-reported or caregiver-reported discrimination. |
| **Intervention** | Development and/or evaluation of measurement tools for assessing discrimination within healthcare settings. | Studies not focused on developing or evaluating measures of discrimination, or not aiming to capture patient or caregiver experiences of discrimination. |
| **Outcomes** | The validity, reliability, or effectiveness of discrimination measurement tools and how they capture patient or caregiver experiences within healthcare. | Review articles, study protocols, pre-prints, concept analysis papers, commentaries, case reports, or expert opinions. Studies focused on discrimination outside of healthcare settings. |
| **Study design** | Original research developing and/or evaluating measurement tools. | Evaluations of the association between discrimination and health outcomes. |

### Information Sources and Search Strategy

The search will be conducted in four electronic databases: MEDLINE via PubMed, Embase via Elsevier, PsycINFO via EBSCOHost, and Web of Science Core Collection via Clarivate. An expert medical librarian will devise and conduct the searches, with input on keywords and final review from the author team. Search strategies will be validated against a set of pre-selected articles and will be peer-reviewed by a librarian using a modified PRESS Checklist.[21] Search terms will include a mix of keywords and subject headings (e.g., MeSH terms) and encompass four key concepts: “perceived discrimination,” “healthcare settings,” “surveys, instruments, questionnaires,” and “development/validation of measurement instruments.” Search hedges or database filters will be used to remove publication types such as editorials, letters, comments, and case reports, and animal-only studies. No limits will be placed on dates. Supplemental search strategies include reviewing the references of included articles and citation tracking.[14] The full reproducible search strategies for all databases will be available in the published manuscript.

All search results will be exported into EndNote and imported into Covidence, a systematic review screening software, and duplicates will be automatically or manually removed as needed.[22]

### Study Selection and Data Extraction

Study selection will be performed in a systematic manner to ensure the identification of all relevant studies that meet the predefined inclusion and exclusion criteria. The process involves several stages: (1) initial screening of identified records by two independent reviewers based on the title and abstract, with retained records undergoing full-text review for eligibility; (2) resolution of conflicts through discussion or by a third reviewer, where only the most comprehensive or recent report will be included for studies reported in multiple publications; (3) creation and maintenance of A Preferred Reporting Items for Systematic Reviews and Meta-Analyses (PRISMA) study inclusion diagram to document the study selection process, including reasons for exclusion at each stage.

For studies meeting the eligibility criteria, data will be dual extracted by two individuals to ensure accuracy. Key data to be extracted includes the year of publication, authors, study locations, research aims, study design, sampling methods, sample size, characteristics of participants, details of healthcare settings, psychometric properties of the measures, and data collection methods for discrimination measurement.

### Quality Assessment (Risk of bias in individual studies)

For quality appraisal of studies we will utilize the COnsensus-based Standards for the selection of health Measurement INstruments (COSMIN) Risk of Bias tool. [23] This tool allows researchers to assess studies systematically and transparently by evaluating key aspects such as the completeness of the research question, including the precise outcome measurement instrument and target construct, as well as whether the study specifies the type of reliability or measurement error assessed (e.g., ICC, Kappa, SEM). It also addresses study design standards, including the stability of subjects over repeated measurements, appropriateness of time intervals, consistency of measurement conditions, and the implementation of blinding procedures. Additionally, it covers the use of preferred statistical methods for different data types, ensuring that appropriate reliability statistics (e.g., ICC for continuous scores, Kappa for ordinal scores) and measurement error parameters (e.g., SEM, LoA) are calculated according to the study design. This approach ensures a thorough assessment of both methodological rigor and the potential impact of design flaws on study reliability.

### Data Synthesis

Narrative synthesis methods will be used to evaluate the measurement tools for discrimination in healthcare settings. Our narrative synthesis will have a two-pronged approach, starting with data synthesis of the psychometric properties of the tools, followed by recommendations based on the results.[24] In the synthesis phase of our systematic review, we will first identify all the key dimensions of measuring discrimination in healthcare. For each dimension, we will provide a concise account of the contributions made by each measurement study. Conflicting findings will be addressed as valuable insights, explained by the differing paradigms from which the data were generated. In the recommendations phase, through reflection, dialogue, and consultation with intended users of the review, we will summarize the key messages from the literature alongside other relevant factors. We will then formulate clear and actionable recommendations for health practice, policy, and further research. No quantitative meta-analysis will be conducted for our results. A meta-analysis is not appropriate for this review because the measurement items we are assessing are designed for different contexts, making direct comparison or aggregation of their results problematic. Instead, we will focus on synthesizing the quality of the individual measures and evaluating their suitability for specific applications. This approach allows for a more nuanced understanding of each measure’s strengths, limitations, and appropriate use cases, which would be lost in a meta-analysis.

### Ethical Considerations

As this systematic review uses secondary data without direct interaction with human subjects, ethical approval is not required by an institutional review board.

### Data Management Plan

No data management plan is necessary given there is no sensitive or personally identifying information that is generated by this study.[25]

## Discussion

Identifying accurate and valid tools to assess discrimination in healthcare is the primary goal of this systematic review, as these measures are vital for addressing health inequities effectively. By identifying and evaluating the psychometric properties of existing measures, this review will help researchers select tools to accurately gauge individual experiences of discrimination or provide insight on the need for new or updated measures. This is particularly important as reliable discrimination measures are foundational for developing interventions aimed at reducing health inequities.

Furthermore, by synthesizing the accuracy of discrimination measurement, this systematic review will support ongoing efforts to identify and mitigate the adverse consequences of discrimination in healthcare, thereby complementing broader initiatives aimed at advancing health equity. A major strength of this review is its comprehensive search strategy, which includes multiple databases and supplemental search techniques, in accordance with National Academies Press systematic review standards. Additionally, the use of the established COSMIN 2020 Risk of Bias tool, ensures a thorough evaluation of the included studies. An established reporting standard, PRISMA-COSMIN for OMIs 2024, provides an additional layer of rigor to our work.

This review will synthesize the existing tools for the measurement of healthcare discrimination. This synthesis should inform future research on the development and validation of comprehensive measurement tools that can capture intersectional experiences of discrimination, including those based on race, ethnicity, gender, and age. Measuring discrimination in healthcare settings presents several challenges, including variation in self-reported experiences and the contextual sensitivity of measurement tools. Different populations may perceive and report discrimination differently, influenced by cultural and socio-economic factors. Addressing these challenges requires the development of culturally sensitive and context-specific measurement instruments. By clarifying the strengths and weaknesses of current tools, this review will provide essential guidance for refining approaches to measuring healthcare discrimination, ensuring that future efforts are both inclusive and effective.

## Data Availability

No data was generated since this is a protocol for an on-going study.

## Acknowledgements

We appreciate the Duke Medical Library for peer-reviewing the search alongside providing resources that guided us.

